# Early prediction of mortality risk among severe COVID-19 patients using machine learning

**DOI:** 10.1101/2020.04.13.20064329

**Authors:** Chuanyu Hu, Zhenqiu Liu, Yanfeng Jiang, Xin Zhang, Oumin Shi, Kelin Xu, Chen Suo, Qin Wang, Yujing Song, Kangkang Yu, Xianhua Mao, Xuefu Wu, Mingshan Wu, Tingting Shi, Wei Jiang, Lina Mu, Damien C Tully, Lei Xu, Li Jin, Shusheng Li, Xuejin Tao, Tiejun Zhang, Xingdong Chen

**Affiliations:** Tongji Hospital, Tongji Medical College, Huazhong University of Science and Technology, Wuhan, Hubei, China; State Key Laboratory of Genetic Engineering, Human Phenome Institute, and School of Life Sciences, Fudan University, Shanghai, 200438, China; Fudan University Taizhou Institute of Health Sciences, Taizhou, China; Key Laboratory of Public Health Safety, Fudan University, Ministry of Education, China and Department of Epidemiology, School of Public Health, Fudan University, Shanghai, China; Health Science Center, Shenzhen Second People’s Hospital. The First Affiliated Hospital of Shenzhen University, Shenzhen, China; Department of Infectious Diseases, Huashan Hospital, Fudan University, Shanghai, China; Department of Biostatistics, School of Public Health, Fudan University, Shanghai, China; Department of Epidemiology and Environmental Health, School of Public Health and Health Professions, University at Buffalo, State University of New York, Buffalo, NY, USA; Department of Infectious Disease Epidemiology, London School of Hygiene & Tropical Medicine, London, UK; Emergency Medicine Department of the Affiliated Hospital of Xuzhou Medical University, Xuzhou, China

**Keywords:** COVID-19, death, fatality rate, predictive model, machine learning

## Abstract

**Background:** Coronavirus disease 2019 (COVID-19) caused by severe acute respiratory syndrome coronavirus 2 (SARS-CoV-2) infection has been spreading globally. The number of deaths has increased with the increase in the number of infected patients. We aimed to develop a clinical model to predict the outcome of severe COVID-19 patients early.

**Methods:** Epidemiological, clinical, and first laboratory findings after admission of 183 severe COVID-19 patients (115 survivors and 68 nonsurvivors) from the Sino-French New City Branch of Tongji Hospital were used to develop the predictive models. Five machine learning approaches (logistic regression, partial least squares regression, elastic net, random forest, and bagged flexible discriminant analysis) were used to select the features and predict the patients’ outcomes. The area under the receiver operating characteristic curve (AUROC) was applied to compare the models’ performance. Sixty-four severe COVID-19 patients from the Optical Valley Branch of Tongji Hospital were used to externally validate the final predictive model.

**Results:** The baseline characteristics and laboratory tests were significantly different between the survivors and nonsurvivors. Four variables (age, high-sensitivity C-reactive protein level, lymphocyte count, and d-dimer level) were selected by all five models. Given the similar performance among the models, the logistic regression model was selected as the final predictive model because of its simplicity and interpretability. The AUROCs of the derivation and external validation sets were 0.895 and 0.881, respectively. The sensitivity and specificity were 0.892 and 0.687 for the derivation set and 0.839 and 0.794 for the validation set, respectively, when using a probability of death of 50% as the cutoff. The individual risk score based on the four selected variables and the corresponding probability of death can serve as indexes to assess the mortality risk of COVID-19 patients. The predictive model is freely available at https://phenomics.fudan.edu.cn/risk_scores/.

**Conclusions:** Age, high-sensitivity C-reactive protein level, lymphocyte count, and d-dimer level of COVID-19 patients at admission are informative for the patients’ outcomes.

## Introduction

The severe acute respiratory syndrome coronavirus 2 (SARS-CoV-2) has emerged in December 2019 and has since spread to nearly 200 countries and territories and has infected more than 0.7 million people by the end of March 2020.^1^ Approximately 36,000 people have died from coronavirus disease 2019 (COVID-19) worldwide.^1^ According to the WHO report, the crude fatality rate of COVID-19 varies from country to country. As of 31 March 2020, the highest fatality rate was observed in Italy (nearly 11%), followed by Spain (nearly 8%) and Iran (nearly 6%).^1^ The fatality rate is also heterogeneous within a country.^2^ In China, the highest rate was found in Wuhan (nearly 5%).^3-5^ In most other provinces, the fatality rate was < 1%.

Epidemiological, clinical, and laboratory features are significantly different between survivors and nonsurvivors.^4,5^ For instance, the nonsurvivors are older than survivors. Dyspnea, chest tightness, and disorder of consciousness are more common in deceased patients than in recovered patients. Concentrations of alanine aminotransferase, aspartate aminotransferase, creatinine, creatine kinase, lactate dehydrogenase, cardiac troponin I, N-terminal pro-brain natriuretic peptide, and d-dimer are markedly higher in nonsurvivors than in survivors.^4,6,7^ Ascertaining the key factors that contribute to the patients’ outcomes is critical for reducing mortality risk and fatality rate. However, these multilevel data may confuse the clinicians with regard to what features indeed impact the COVID-19 patients’ outcomes. In the current study, we aimed to develop a clinical model to predict the mortality risk of severe COVID-19 patients based on epidemiological, clinical, and the first laboratory test data after admission.

## Methods

### Study participants and covariate collection

In total, 256 severe laboratory-confirmed COVID-19 patients (126 survivors and 130 nonsurvivors) admitted to the Sino-French New City Branch of Tongji Hospital, Wuhan, between 28 January 2020 and 11 March 2020 were included. Tongji Hospital was urgently rebuilt and has been assigned by the Chinese government as a designated hospital for severely or critically ill patients with COVID-19.^4^ We collected the epidemiological (e.g., age and sex), clinical (e.g., fever or not and computed tomography [CT] imaging features) and the first laboratory data after admission (e.g., the high-sensitivity C-reactive protein [hsCRP] level) from the patient’s medical records (Figure S1). For further details see Table S1. The values of biochemistry indexes tested more than three days after admission were excluded even if they were part of the first test. For values that were left or right truncated (e.g., d-dimer > 21 µg/mL), we used the values at the truncated point as the surrogates (e.g., using 21 µg/mL for those with d-dimer level > 21 µg/mL). As shown in Figure S2, patients who were with >10% missing values, stayed in the hospital < 7 days, afflicted by a severe disease before admission (e.g., cancer, aplastic anemia, and uremia), were unconscious at admission, and were directly admitted to the intensive care unit (ICU) were excluded. Finally, 183 patients were included to construct the predictive models; among them, 115 recovered and were discharged, and 68 were died from COVID-19. This study was approved by the National Health Commission of China and Ethics Commission of Tongji Hospital (TJ-IRB20200402). Written informed consent was waived by the Ethics Commission of the designated hospital for emerging infectious diseases.^8^

## Model development

The development of the predictive model consisted of three main stages: 1) data preprocessing; 2) variable selection and model evaluation; and 3) external validation (Figure 1).

**Figure 1.**
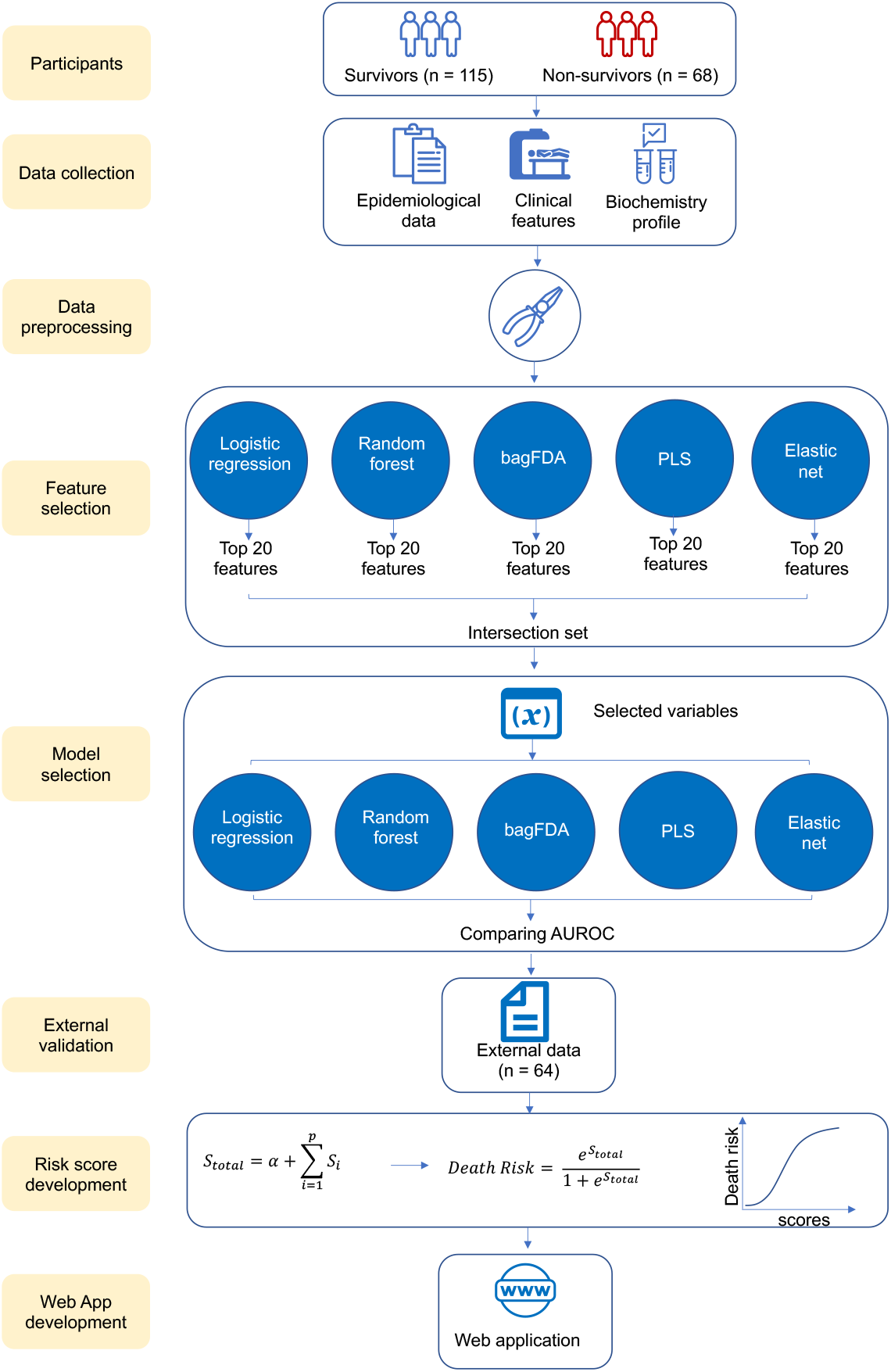
The study flow chart. (Abbreviations: bagFDA, bagged flexible discriminant analysis; PLS, partial least squares; AUROC, area under the receiver operating characteristic curve)

### Data preprocessing

Covariates with >30% missing data were excluded. For pairs of highly correlated variables (correlation coefficient >0.9), we removed the variable with the higher missing rate. Variable with zero or near-zero variance were also excluded. A rule of thumb for detecting predictors with zero or near-zero variance is as follows: 1) the number of unique values divided by the sample size is small (set to 10% in our study); and 2) the ratio of the frequency of the most prevalent value to the frequency of the second most prevalent value is large (set to 20 here). Non-normally distributed continuous variables were transformed using a Box-Cox transformation. Missing values were imputed using bagging trees. In total, 51 covariates were finally included.

### Variable selection and model evaluation

Considering the potential linear and curvilinear relationships between the predictors and the outcome, we selected five machine learning models (logistic regression, partial least squares [PLS] regression, elastic net model, random forest, and bagged flexible discriminant analysis [FDA]) to fit the data. Ten-fold cross validation and the areas under the receiver operating characteristic curves (AUROCs) were used to measure the models’ performance. The selected tuning parameters and performance of the models based on the full data are shown in Table S2. For logistic regression, we used stepwise backward to select the variables. The variable importance was assessed using the absolute value of the t-statistic. For PLS regression, the variable importance measure here was based on the weighted sums of the absolute regression coefficients. The weights were a function of the reduction in the sums of squares across the number of PLS components and were computed separately for each outcome. Therefore, the contribution of a coefficient was weighted proportionally to the reduction in the sums of squares. For the elastic net model, the selected variables were those of coefficients that did not shrink to 0. For the random forest model, the prediction accuracy for the out-of-bag portion of the data was recorded for each tree. Then, the same procedure was performed after permuting each predictor variable. The difference between the two accuracies was then averaged across all the trees and normalized by the standard error.^9^ For the bagged FDA model, a series of cutoffs were applied to the predictor data to predict the class. The AUROC, sensitivity, and specificity were computed for each cutoff. The trapezoidal rule was used to compute the AUROC, which was used as the measure of variable importance.^10^ The top 20 most important variables selected by the five models are shown in Figure 2A-E. We chose the intersection set of these variables. Four variables were finally selected (age, hsCRP, lymphocyte count, and d-dimer). The five models were refitted using these four variables. The AUROC, sensitivity, and specificity, obtained from 10-fold cross validation were used to evaluate the models’ performance.

**Figure 2.**
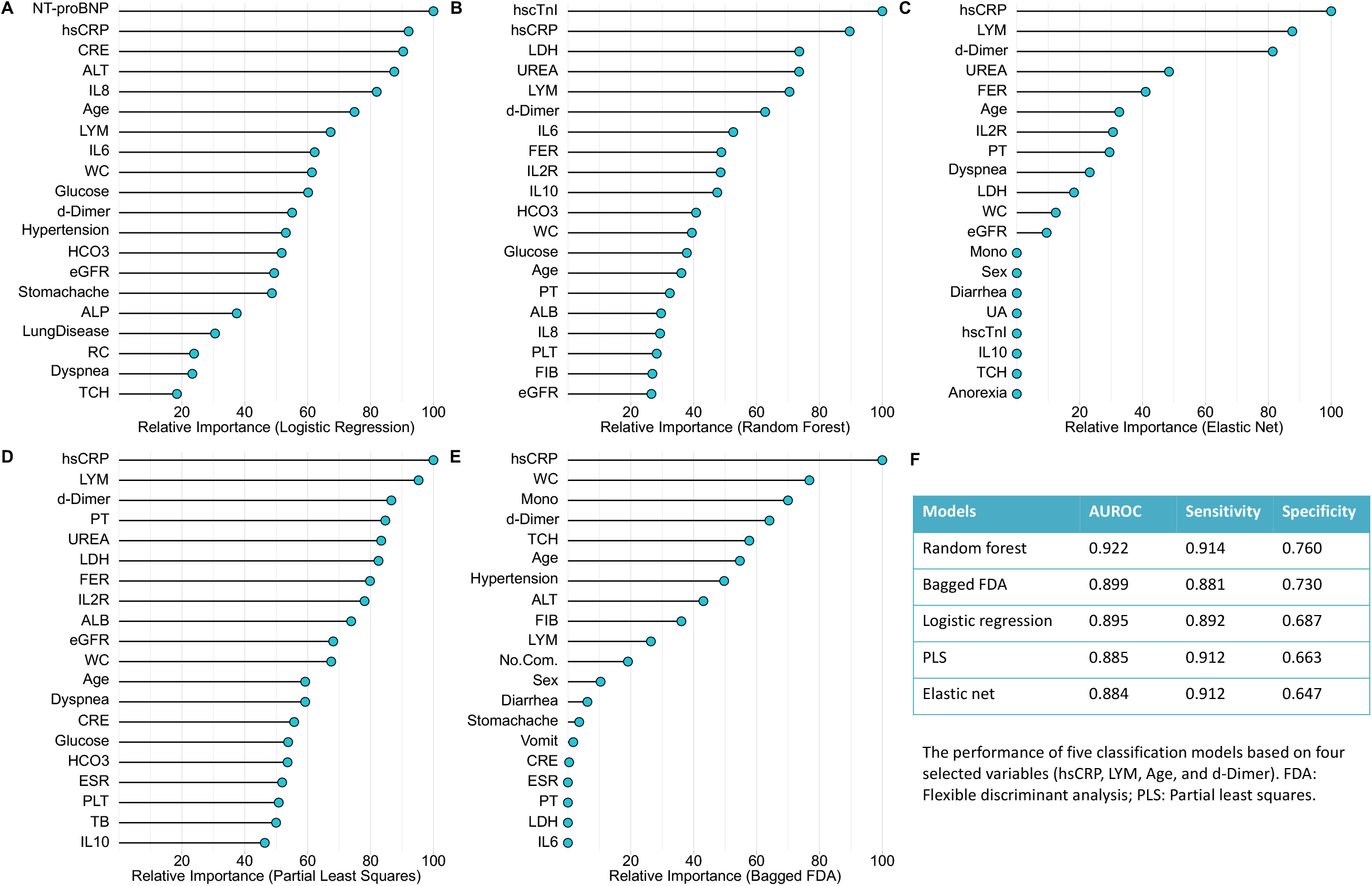
The top 20 important variables selected by five machine learning models (A-E) and the model performance based on the selected variables (F). (Abbreviations: NT-proBNP, N-terminal pro-brain natriuretic peptide; hsCRP, high-sensitivity C-reactive protein; CRE, creatinine; ALT, alanine aminotransferase; IL8, interleukin 8; LYM, lymphocyte count; IL6, interleukin 6; WC, white cell count; eGFR, estimated glomerular filtration rate; ALP, alkaline phosphatase; RC, red cell count; TCH, total cholesterol; FIB, fibrinogen; PLT, platelet count; TB, total bilirubin; ALB, albumin; PT, prothrombin time; IL10, interleukin 10; IL2R, interleukin-2 receptor; FER, ferritin; LDH, lactate dehydrogenase; hscTnI, high-sensitivity cardiac troponin I; Mono, monocyte count; UA, uric acid; ESR, erythrocyte sedimentation rate; No.Com., number of basic conditions)

### External validation

In total, 64 severe laboratory-confirmed COVID-19 patients (33 survivors and 31 nonsurvivors) admitted to the Optical Valley Branch of Tongji Hospital, Wuhan, were included as the external validation set. We used the selected predictive model to predict the probability of death in these patients. The AUROC, sensitivity, and specificity were used to evaluate the model performance.

### Cost curves of the predictive model

Cost curve is a graphical technique for visualizing the performance (expected cost) of 2-class classifiers over a range of possible class distributions and cutoffs. Herein, we defined the cost of false negative was three times more than that of false positive. The cutoffs predicting the death were ranged from 0.1 to 0.9. The false negative rates and false positive rates were calculated by the confusion matrix of the predictive model using different cutoffs. We then visualized the curves between the normalized true positive rates and the normalized expected costs.^11^ The normalized true positive rates can be calculated as: 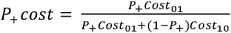 and the normalized expected costs can be calculated as: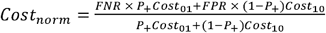, where the *P*_+_ denotes the true positive rate, *Cost*_01_ denotes the cost of false negative, *Cost*_10_ denotes the cost of false positive, FNR denotes the false negative rate, and FPR denotes the false positive rate.^11^

### Statistical analysis

Continuous and categorical variables are presented as means (standard deviations) (or medians (interquartile range)) and frequencies (percentages), respectively. We used Student’s t-tests, Mann-Whitney U tests, χ^2^ tests, and Fisher’s exact tests, where appropriate, to compare the differences between survivors and nonsurvivors. All analyses were implemented in R 3.6.1 (R Foundation for Statistical Computing, Vienna, Austria). The model development and validation were implemented using the *caret* package (version 6.0-85).^12^

## Results

### Demographics and baseline characteristics of survivors and nonsurvivors

The covariates included in the predictive models are shown in Table 1. In general, most characteristics were significantly different between the survivors and nonsurvivors. The nonsurvivors were more likely to be male and older than the survivors. The proportions of basic conditions (e.g., diabetes and hypertension) were comparable between these two samples. For symptoms at disease onset, the nonsurvivors reported loss of appetite, dyspnea, and productive cough more often than the survivors. The laboratory test indexes were also significantly different between the deceased patients and those who recovered. For example, the white cell count was higher in the nonsurvivors than the survivors. Conversely, the lymphocyte count was nearly twice as high in the survivors than in the nonsurvivors. The nonsurvivors had more liver and kidney function injuries. Specifically, the concentrations of liver enzymes, urea, and creatinine were markedly higher in the nonsurvivors, whereas the albumin level and estimated glomerular filtration rate were lower in this sample. The levels of all the inflammatory factors were higher in the nonsurvivors than in the survivors. The circulating levels of hsCRP and d-dimer were more than 6-fold and nearly 3-fold higher in nonsurvivors than in survivors, respectively.

**Table 1.**
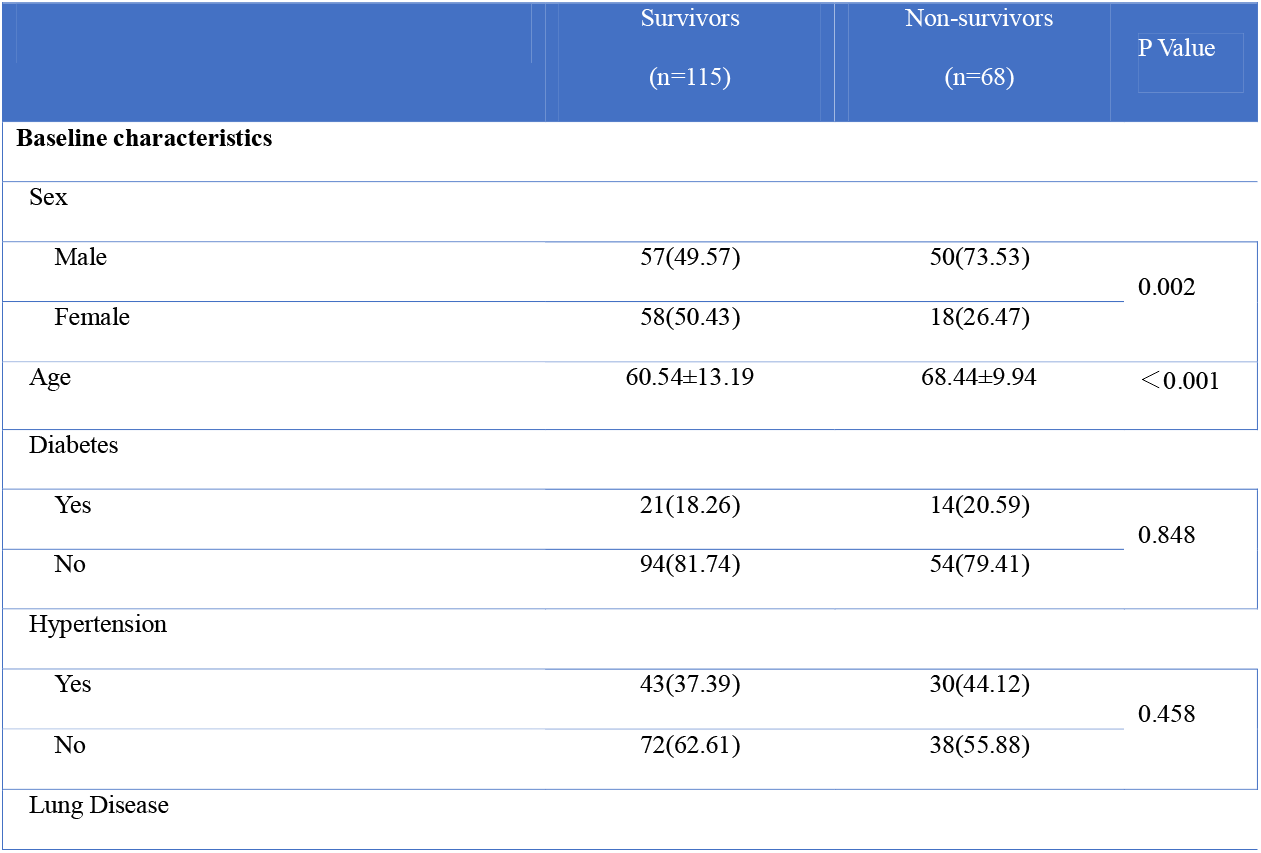
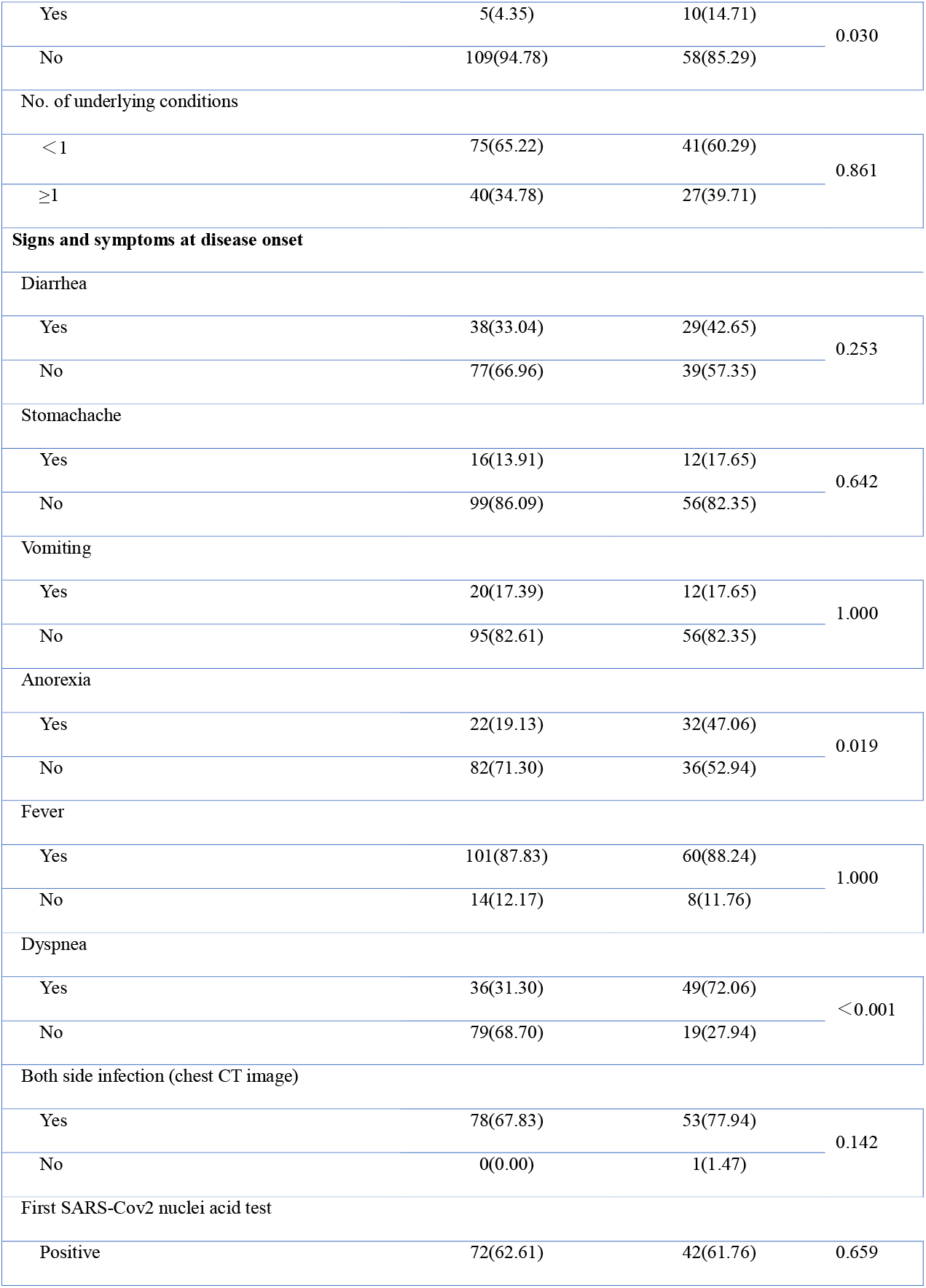
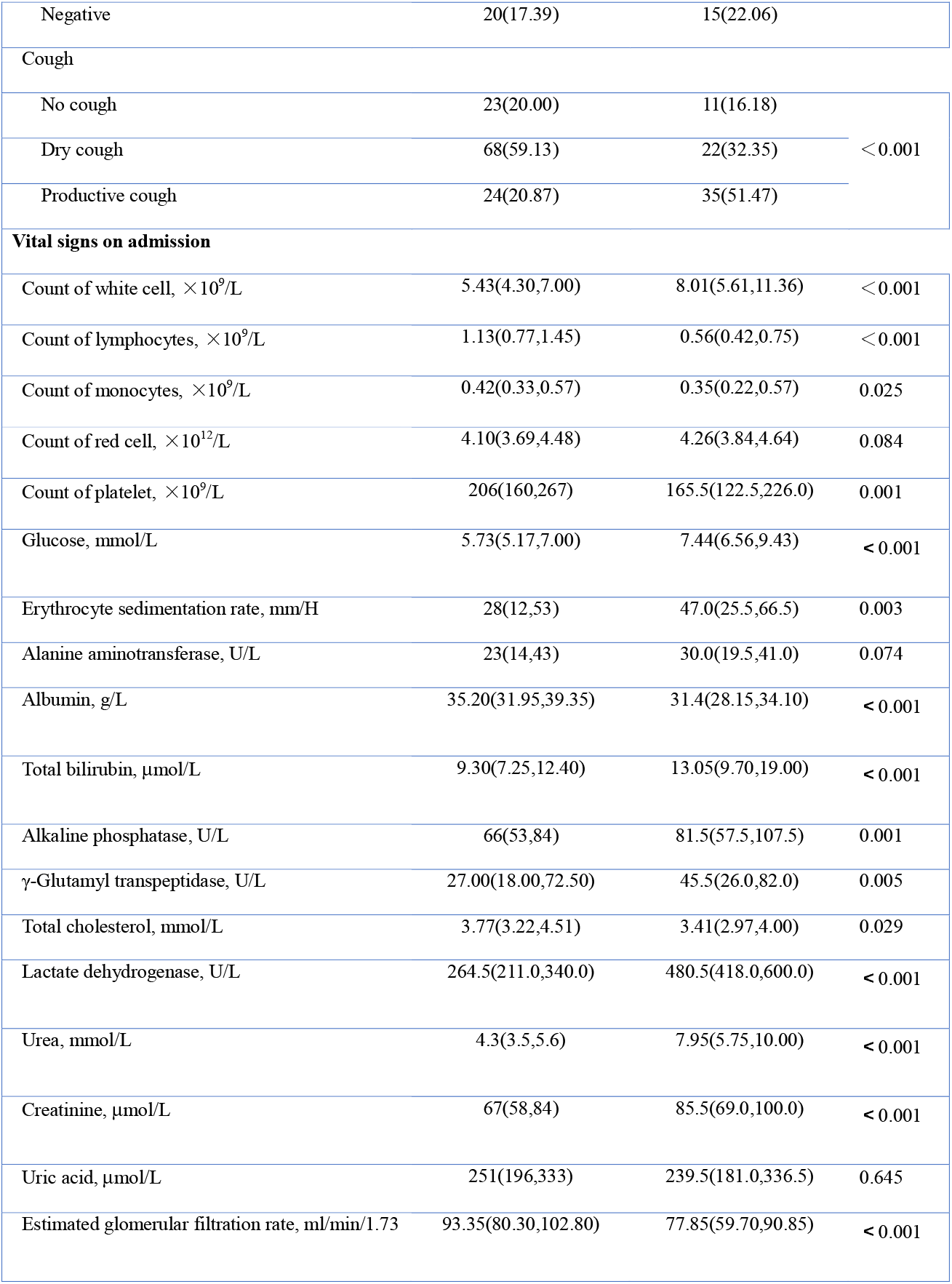
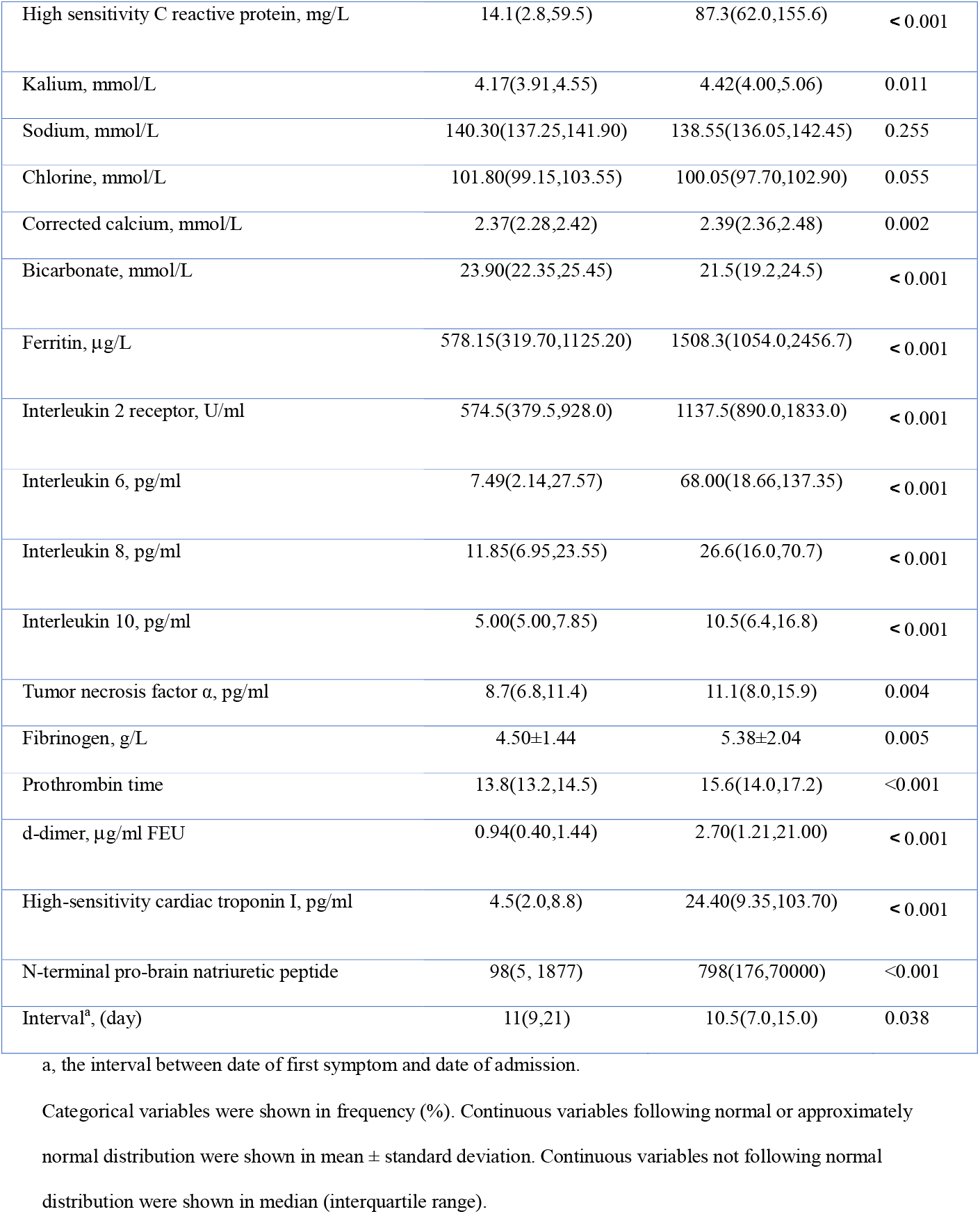
The baseline characteristics and laboratory findings at admission included in the predictive models.

### Model development and External validation

The process of model development is detailed in the Method section and in Figure 1. The AUROC obtained from 10-fold cross validation was used to compare the performance of five selected predictive models (Figure 2F). Considering the minor differences between the AUROC of the logistic regression model (0.895) and that of the random forest (0.922) and bagged FDA (0.899), we selected the logistic regression model as the final model because of its simplicity and high interpretability. The AUROCs of the logistic regression model based on age, hsCRP, lymphocyte count, and d-dimer, in combination and individually, are shown in Table S3. The model performance significantly increased when using the four variables in combination (Figure 3A). The AUROC of the external validation set was 0.881, with a sensitivity of 0.839 and specificity of 0.794 for predicting death when using a probability of death of 50% as the cutoff (Figure 3B).

**Figure 3.**
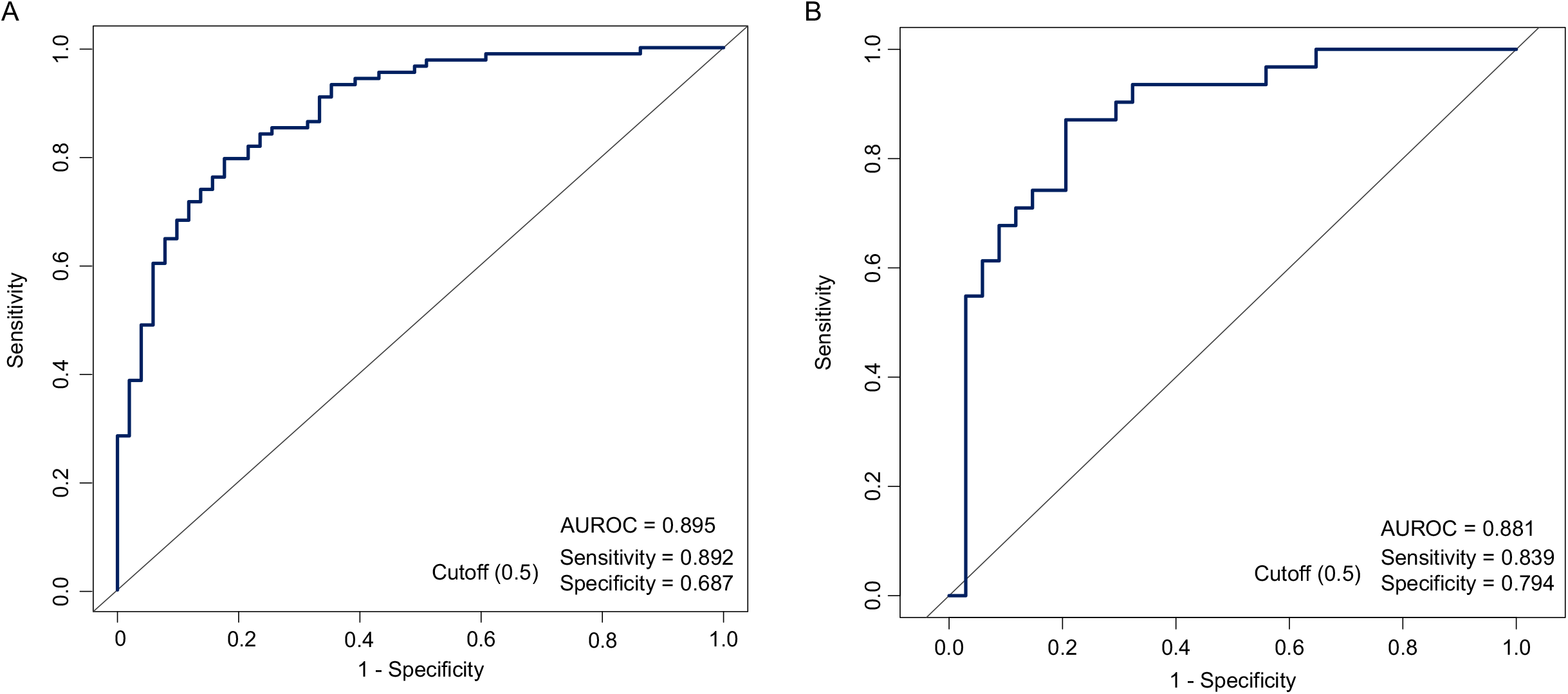
The area under the receiver operating characteristic curve (AUROC) of the logistic regression model based on selected variables in the derivation set (A) and the external validation set (B).

### Cost curves of the predictive model

As shown in Figure 4A, the death probability was curvilinearly associated with the normalized *P*_+_. Figure 4B showed the cost curves with the cutoff ranged from 0.1 to 0.9. When the death probability is <10% (i.e., normalized *P*_+_ <0.25), the lowest expected cost was observed using cutoff of 0.8. When the death probability increases to 25% (i.e., normalized *P*_+_ = 0.5), the lowest expected cost was observed with cutoff of 0.6. When the death probability reaches at nearly 50% (i.e., normalized *P*_+_ around at 0.75), the lowest expected cost was observed with cutoff of 0.2.

**Figure 4.**
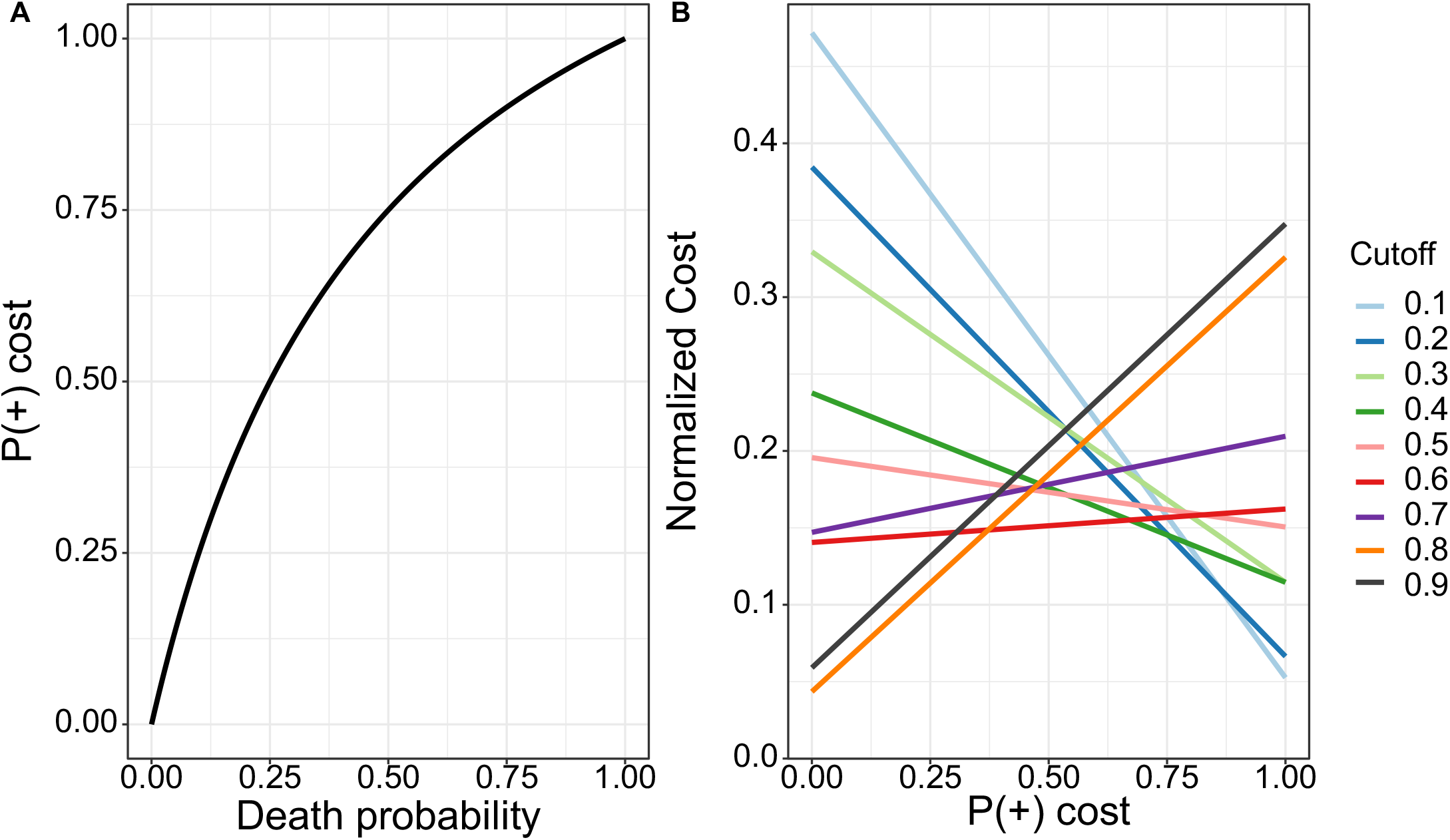
The relationship between probability of death and the normalized probability (A) and the cost curves of the predictive models using different cutoffs (B).

### Development of risk scores and a web application

The contributions of the selected variables were assessed using the logistic regression model. The regression coefficients were calculated as the weight of each predictor. The risk score was therefore calculated as: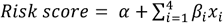, where *α, x*_*i*_, and *β* _*i*_ denote the regression intercept, observed value and coefficients, respectively, of the *i*^th^ predictor (Figure 5A). The probability of death can be calculated via 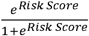 Figure 5B shows that the risk scores in both the survivors and nonsurvivors are approximately normally distributed. A S-shaped correlation pattern between the risk scores and probability of death is shown in Figure 5C. The mortality risk exceeds 50% when the risk score is >0.

**Figure 5.**
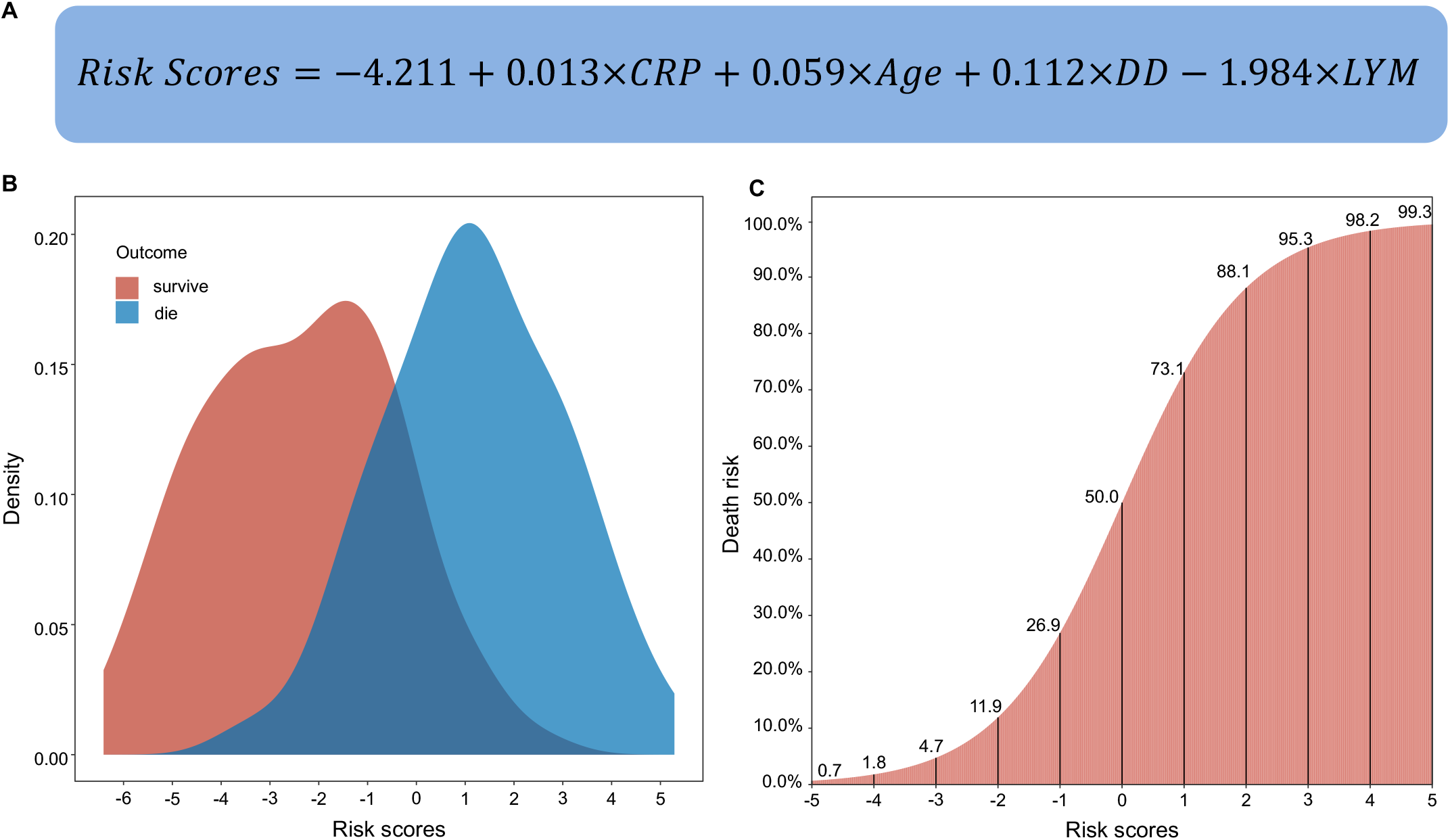
The formula to calculate the risk scores (A) and their distributions among survivors and nonsurvivors (B) and the corresponding probability of death (C).

To facilitate the application of our predictive model, we also developed an accompanying web tool (https://phenomics.fudan.edu.cn/risk_scores/). Readers can freely access this website and input the values of hsCRP, age, lymphocyte count, and d-dimer to predict the mortality risk and its 95% confidence interval of a COVID-19 patient.

## Discussion

COVID-19 is currently a worldwide pandemic.^13-15^ The number of laboratory-confirmed patients as well as the number of related deaths are continuously increasing. The fatality rate might further increase along with the increasing number of infected people, because even the most advanced healthcare systems are likely to be overwhelmed.^16,17^ The Chinese Centers for Disease Control and Prevention recently reported that out of more than 70,000 confirmed cases, most of them were classified as mild or moderate, and approximately 20% were classified as severe or critical.^18^ Even if we assume that the fatality rate is 10% in severe and critically ill cases,^4^ the number of COVID-19 induced deaths is considerable because of the enormous number of infected patients. Identifying the patients at high risk of death is critical for patient management and to reduce the fatality rate. In this clinical prediction modeling study, we took full advantage of the multifaceted data of COVID-19 patients at admission to predict their outcomes. Four variables (i.e., age, hsCRP, d-dimer, and lymphocyte count) were selected and used to fit a logistic regression model. The predictive performance of our model was acceptable in both the derivation set and the external validation set. We also developed a web tool to implement our predictive model. Clinicians can use this web tool to predict the mortality risk of COVID-19 patients early. For those patients with a relatively higher probability of death (e.g., >40%), more interventions could be adopted at an earlier stage by clinicians to reduce the mortality risk.

In a recent study, older age, d-dimer levels greater than 1 μg/mL, and a higher sequential organ failure assessment (SOFA) score at admission were reported to be associated with higher odds of in-hospital death.^3^ The fatality rate was highly heterogeneous among patients of different ages. For instance, the overall COVID-19 case fatality rate in China was estimated as 0.32% in those aged <60 years old and substantially increased to 6.4% in those aged ≥60 years old.^19^ Among those aged 80 years old and older, this rate was as high as 13.4%.^19^ Likewise, in Italy, the fatality rate increased from 0.3% among patients aged 30-39 years old to 20.2% among those aged ≥80 years old.^20^ In 2019, approximately 23% of the Italian population was aged 65 years or older. This percentage may explain, in part, Italy’s higher case-fatality rate compared with that of other countries.^20^ In our study, age was selected as a key factor in all the predictive models. The age-dependent deterioration in immunological competence (e.g., B-cell and T-cell deficiencies), often referred to as “immunosenescence”, and the excess production of type 2 cytokines could lead to a deficiency in the control of viral replication and more prolonged proinflammatory responses, potentially leading to a poor outcome.^21,22^ In this study, the deceased patients had persistent and more severe lymphopenia compared with recovered patients and the lymphocyte count was selected and incorporated into the predictive model. Defects in function of lymphocytes are age-dependent and are associated with inflammation levels.^23,24^

Additionally, hsCRP, the most commonly used inflammatory biomarker in the clinic, was selected for our predictive models. Patients with higher levels of hsCRP at admission were deemed to have higher levels of inflammation. In our study, we found that the hsCRP level was more than 6 times higher in the nonsurvivors than in the survivors. Although the levels of other inflammatory biomarkers, such as ferritin and the erythrocyte sedimentation rate, were also elevated in the nonsurvivors, they were not selected and incorporated in the predictive model. This omission might be explained by the higher missing rates of these covariates and their high correlation with hsCRP. Coagulation disorder, characterized by an elevated prothrombin time and d-dimer level, was also frequently observed among COVID-19 patients.^25,26^ In our study, the prothrombin time and d-dimer were significantly higher in the non-survivors than in the survivors. These higher values suggest an increased risk of disseminated intravascular coagulation, which was one of the frequently diagnosed complications among the later stage of the COVID-19.^4,25,27^

Our study has a number of strengths. First, to our knowledge, this is the first study for predicting the outcomes of severe COVID-19 patients based on comprehensive data. Second, to ensure the robustness of our predictive model, we enforced strict inclusion and exclusion criteria on the included participants and study data. Third, we used advanced modeling strategies to select features and to construct the predictive models. The final model is simple (it included only four variables) and highly interpretable (the model is a linear model, and the effects of the predictors are reflected by the regression coefficients). Moreover, we externally validated the final predictive model. Fourth, we developed an accompanying web tool to facilitate the application of our predictive model by clinicians. Our study also has limitations. First, the predictive models were constructed based on a relatively small sample size; the interpretation of our findings might be limited. Second, due to the retrospective study design, not all the laboratory tests were performed in all the patients. Some of them might be deleted in the data preprocessing procedure and their roles might be underestimated in predicting patients’ outcomes. Third, patients were sometimes transferred from other hospitals to the two branches of Tongji hospitals, although we excluded patients who did not meet the inclusion criteria. The values of the laboratory tests might be biased by prior antiviral treatment in these patients. Finally, the patients in the derivation set and the validation set were from Tongji Hospital, which is one of the hospitals with a high level of medical care in China. Some critically ill patients recovered here might die in other hospitals with suboptimal or typical levels of medical care. The cutoff for predicting death should be <50% (e.g., defining patients who have a >30% probability of death as high-risk patients) in these settings.

In summary, using available clinical data, we developed a robust machine learning model to predict the outcome of COVID-19 patients early. Our model and the accompanying web application are of importance for clinicians to identify patients at high risk of death and are therefore critical for the prevention and control of COVID-19.

## Data Availability

All data were collected from medical records in Tongji Hospital, Wuhan, China.

## Contributors

ZL, TZ, XC, and CH conceived the study design. CH, QW, SL, XT, XL, and YS collected the data. ZL, YJ, XZ, KX, and OS performed the statistical analysis. ZL, CH, YJ, XZ, OS, KY, and XC wrote the manuscript. All authors provided critical revisions of the draft and approved the submitted draft. The corresponding author attests that all listed authors meet authorship criteria and that no others meeting the criteria have been omitted. XC is the guarantor.

